# Diagnostic accuracy of SARS-CoV-2 rapid antigen detection testing in symptomatic and asymptomatic children in the clinical setting

**DOI:** 10.1101/2021.04.15.21255577

**Authors:** Arnaud G. L’Huillier, Matthieu Lacour, Debora Sadiku, Mehdi A. Gadiri, Loraine De Siebenthal, Manuel Schibler, Isabella Eckerle, Selina Pinösch, Laurent Kaiser, Alain Gervaix, Alban Glangetas, Annick Galetto-Lacour, Laurence Lacroix

## Abstract

**Importance:** Antigen-based rapid diagnostic tests (RDTs) have shown good sensitivity for SARS-CoV-2 detection in adults and are used in children despite the lack data from children.

**Objective:** We evaluated the diagnostic performance of the Panbio™-COVID-19 Ag Rapid Test Device (P-RDT) in symptomatic and asymptomatic children against reverse-transcription polymerase chain reaction (RT-PCR) on nasopharyngeal swabs (NPS).

**Design:** Prospective diagnostic study from 11.2020 to 03.2021.

**Setting:** Single-center.

**Participants:** Consecutive symptomatic and asymptomatic participants 0-16yo.

**Intervention:** Two NPS for both RT-PCR and P-RDT.

**Main outcome:** P-RDT sensitivity and specificity.

**Results:** Eight-hundred and twenty-two participants completed the study, of which 533 (64.9%) were symptomatic. Among the 119 (14.5%) RT-PCR positive patients, the overall P-RDT sensitivity was 0.66 (95%CI 0.57-0.74). Mean viral load (VL) was higher among P-RDT positive than negative ones (p<0.001). Sensitivity was 0.87 in specimens with VL>1.0E6 copies/mL (95%CI 0.87-1.00), which is the accepted cut-off for the presence of infectious virus, and decreased to 0.67 (95%CI 0.59-0.76) for specimens >1.0E3 copies/mL.

Among symptomatic participants, the P-RDT displayed a sensitivity of 0.73 (95%CI 0.64-0.82), which peaked at 1.00 at 2 days post onset of symptoms (DPOS; 95%CI 1.00-1.00), then decreased to 0.56 (95%CI 0.23-0.88) at 5 DPOS. There was a trend towards lower P-RDT sensitivity in symptomatic children <12 years (0.62 [95%CI 0.45-0.78]) versus ≥12 years (0.80 [95%CI 0.69-0.91]; p=0.09). VL which was significantly lower in asymptomatic participants than in symptomatic ones (p<0.001). The P-RDT displayed a sensitivity of 0.43 (95%CI 0.26-0.61).

Specificity was 1.00 in symptomatic and asymptomatic children (95%CI 0.99-1.00).

**Conclusion and relevance:** The overall respective 73% and 43% sensitivities of P-RDT in symptomatic and asymptomatic children was below the 80% cut-off recommended by the World Health Organization. These findings are likely explained by lower VLs in children at the time of diagnosis. As expected, we observed a direct correlation between VL and P-RDT sensitivity as well as variation of sensitivity according to DPOS, a major determinant of VL. These data highlight the limitations of RDTs both in symptomatic and asymptomatic children, with the potential exception in early symptomatic children ≥12yrs where sensitivity reached 80%.

## INTRODUCTION

The current Coronavirus Disease 19 (COVID-19) pandemic induces the need for widespread SARS-CoV-2 testing to control virus circulation. The rapid identification of SARS-CoV-2 infected individuals is important whether persons are symptomatic or not, as the role of asymptomatic persons in SARS-CoV-2 transmission is still unclear. Therefore, easy to use, affordable and rapid diagnostic methods are required in addition to the gold-standard reverse-transcription polymerase chain reaction (RT-PCR)^1^. These devices are increasingly helpful in settings where results are immediately needed, access to a testing facility is limited or in case of shortage of RT-PCR reagents. Several antigen-based rapid diagnostic tests (RDTs) have been marketed to fill this gap. Among them, the Panbio™-COVID-19 Ag Rapid Test Device COVID-19 (referred hereby as P-RDT) has displayed an overall sensitivity ranging between 61%-92%^2-7^ in adults when compared to nasopharyngeal RT-PCR. Sensitivity was improved for higher viral loads (VLs), shorter duration of symptoms and/or in symptomatic patients^2-6^. In symptomatic children, the overall sensitivity of the P-RDT was 45-78%^5,8,9^ but published data do not take into account the effect of VL nor the duration of symptoms. Moreover, to our knowledge no study has evaluated this assay in asymptomatic children at time of sampling. The aim of the present study was to provide an independent evaluation of the diagnostic performance of the P-RDT in a large cohort of symptomatic and asymptomatic children. We also aimed to identify situations with optimal P-RDT sensitivity, by accounting for VL, day post onset of symptoms (DPOS) and type of symptoms.

## METHODS

### Setting

This single-center prospective diagnostic study was performed in Geneva University Hospitals’ (HUG) pediatric testing center, from November 10^th^ 2020 to March 26^th^ 2021, with a peak incidence of 583/100’000/week^10^. Participants 0 to 16 years old who presented with the need for SARS-CoV-2 RT-PCR testing were approached. Indication for RT-PCR testing in symptomatic participants was symptoms suggestive of SARS-CoV-2 infection according to local governmental testing criteria. Indications for RT-PCR testing in asymptomatic participants were notification by local health authorities after contact with a laboratory confirmed SARS-CoV-2 infected person and pre-travel testing.

### Study procedures

For each enrolled participant, two nasopharyngeal swabs (NPS) were collected. Nurses were trained to perform NPS testing through a standardized video-documented procedure. First, a standard flocked swab placed in viral transport media (VTM) was used for viral genome detection by RT-PCR. The second swab, provided in the P-RDT kit, was obtained from the contralateral or ipsilateral nostril and was performed immediately at the testing center as per the manufacturer’s instructions. All study participants and/or caregivers provided written informed consent prior to specimen collection. The study was approved by the local research ethics board (Commission cantonale d’éthique de la recherche #2020-02323).

### P-RDT testing

The Panbio™-COVID-19 Ag Rapid Test Device (Abbott Rapid Diagnostics, US) was chosen for the current study based on adult data from our institution showing optimal sensitivity and ease of use. The P-RDT was used as recommended by the manufacturers, using materials provided in the kit only. P-RDT results were read independently by two members of the study team, both being blinded to the result assigned by their pair as well as to the clinical presentation of the participant. Any discrepant result was considered positive when any of the above-mentioned reader set a positive diagnosis.

### RT-PCR testing

Gold standard RT-PCR testing was performed either on Cobas® SARS-CoV-2 assay (cobas® SARS-CoV-2 Test, Cobas 6800, Roche, Switzerland) or on Nimbus RT-PCR assay, using NPS in 3 mL VTM. VL were expressed as SARS-CoV-2 genome copy numbers per mL. A standard curve was obtained by using a quantified supernatant from a cell culture isolate of SARS-CoV-2. All VLs were calculated from the Cycle threshold (Ct)-values, according to log10 SARS-CoV-2 RNA copies/mL = (Ct-44.5)/-3.3372 for Cobas^11,12^ and (Ct-45.92)/-3.488 for Nimbus. Calculations were corrected for input and extraction volumes.

### Data collection

The following data collected at enrolment were managed using RedCap™ managed using REDCap electronic data capture tools hosted at HUG: date of enrolment, number of days post onset of symptoms (DPOS), gender, age, type of symptoms (nasal discharge, cough, dyspnea, dysphagia, dysgueusia, anosmia, vomiting, diarrhea, fever, chills, decreased intake, headache, myalgia, fatigue and irritability, abdominal pain, nausea) and comorbidities if present (chronic respiratory disease, cardiopathy, immunosuppression, cancer, diabetes, obesity, hypertension and organ failure). P-RDT results, RT-PCR and VL/Ct values were included subsequently.

### Statistics

Before study onset, a sample size was calculated to have sufficient power to generate a 95% confidence interval (CI) with a lower bound above the World Health Organization’s (WHO) target of 80%, if the prevalence was 25% (corresponding the pediatric positivity rate at study onset) and the measured sensitivity of 85% (corresponding to the sensitivity reported in adults in our institution)^6^. The sample size was therefore estimated at 654 participants. Continuous variables were expressed by their mean ±standard deviation (SD) and median (interquartile range [IQR]) upon variable distribution. Categorical variables were presented by their frequencies and relative proportions. For comparisons of continuous variables, parametric Student T-tests and nonparametric Mann-Whitney tests upon variable distribution were used. For categorical variables, either Chi^2^ or Fischer’s exact tests were performed depending on applicability. Statistical analyses were processed under SPSS software v23.0 (IBM Corp., Armonk, NY). Statistical significance was defined as p<0.05 (two-sided).

## RESULTS

Eight hundred and eighty-five pediatric participants were enrolled. Among them, 63 were subsequently excluded (eFigure 1 in the Supplement). A total of 822 participants completed the study and had both RT-PCR and P-RDT performed. Demographics of symptomatic and asymptomatic study participants are detailed in Table 1.

**Table 1.** Study participants’ demographics.

| **Characteristics** | **Symptomatic**<br>**(n=533)** | **Asymptomatic**<br>**(n=289)** | **Combined**<br>**(n=822)** | **p-**<br>**value** |
| --- | --- | --- | --- | --- |
| Median age (±IQR) | 12.1 (9.4-14.5) | 10.9 (8.5-13.7) | 11.8 (9.0-14.3) | 0.002 |
| Female sex, n (%) | 266 (49.9) | 138 (47.8) | 404 (49.1) | 0.555 |
| *Comorbidities, n (%)* |  |  |  |  |
| *Chronic respiratory disease* | 33 (6.2) | 13 (4.5) | 46 (5.6) |  |
| *Obesity* | 15 (2.8) | 10 (3.5) | 25 (3.0) |  |
| *Diabetes* | 1(0.2) | 3 (1.0) | 4 (0.5) |  |
| *Hypertension* | 2 (0.4) | 0 | 2 (0.2) |  |
| *Cancer* | - | 2 (0.7) | 2 (0.2) |  |
| *Cardiopathy* | 1 (0.2) | - | 1 (0.1) |  |
| *Other immunosuppression* | 1 (0.2) | - | 1 (0.1) |  |
| *Chronic liver failure* | 1 (0.2) | - | 1 (0.1) |  |
| Result of RT-PCR, n (%) |  |  |  | 0.014 |
| *Negative* | 444 (83.3) | 259 (89.6) | 703 (85.5) |  |
| *Positive* | 89 (16.7) | 30 (10.4) | 119 (14.5) |  |
| Mean log RNA copies/mL (±SD) | 6.6 (±1.6) | 5.1 (±1.7) | 6.2 (±1.8) | <0.001 |

RT-PCR: reverse transcription polymerase chain reaction; RNA: ribonucleic acid; IQR: interquartile range; SD: standard deviation

Overall, 14.5% (119/822) were positive by RT-PCR with a mean RNA VL of log 6.2 copies/mL (SD 1.8) (Table 1). Among the 822 P-RDT performed, only one P-RDT result displayed a discrepant interpretation between the two observers (κ=0.999). The corresponding patient was subsequently considered as positive for the purpose of the analysis, leaving an overall positivity rate of 9.6% (79/822). The P-RDT’s sensitivity and specificity when challenged against RT-PCR were 0.66 (95%CI 0.57-0.74) and 1.00 (95%CI 1.00-1.00) respectively (Table 2 and eTable 1 in the Supplement). Mean VL was higher among positive P-RDT specimens than negative ones (7.1 [SD 1.3] vs 4.6 [SD 1.5]; p<0.001) (Figure 1).

**Table 2.**
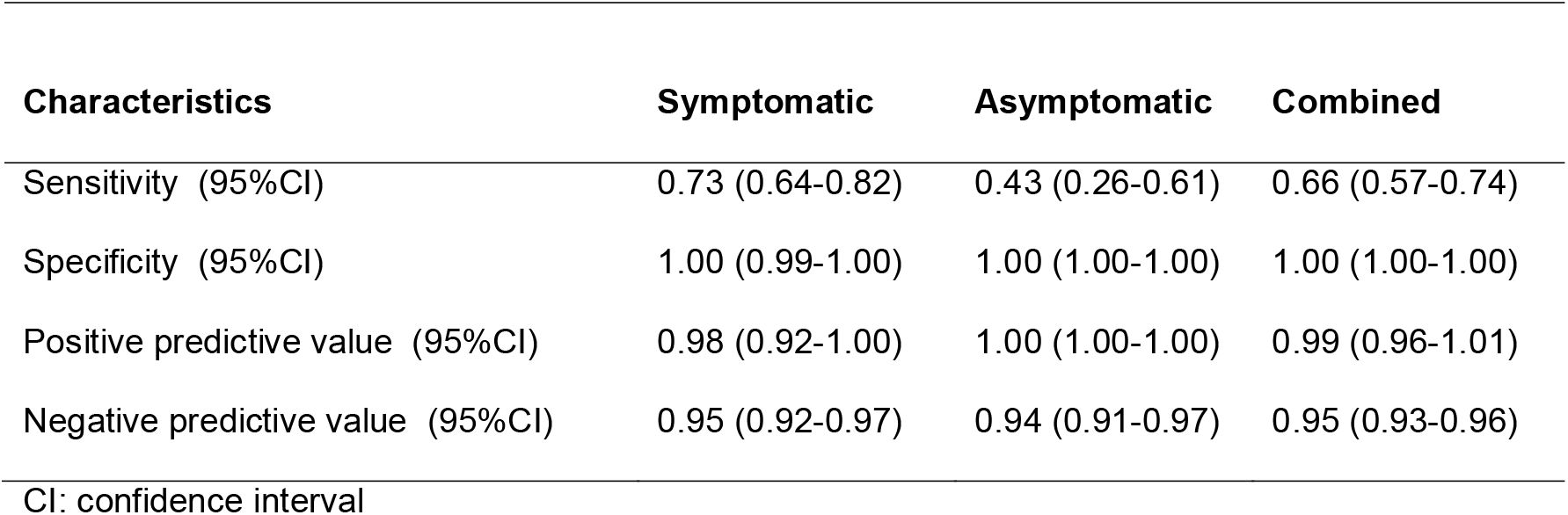
Diagnostic accuracy of the Panbio™ RDT.

| **Characteristics** | **Symptomatic** | **Asymptomatic** | **Combined** |
| --- | --- | --- | --- |
| Sensitivity (95%CI) | 0.73 (0.64-0.82) | 0.43 (0.26-0.61) | 0.66 (0.57-0.74) |
| Specificity (95%CI) | 1.00 (0.99-1.00) | 1.00 (1.00-1.00) | 1.00 (1.00-1.00) |
| Positive predictive value (95%CI) | 0.98 (0.92-1.00) | 1.00 (1.00-1.00) | 0.99 (0.96-1.01) |
| Negative predictive value (95%CI) | 0.95 (0.92-0.97) | 0.94 (0.91-0.97) | 0.95 (0.93-0.96) |
| CI: confidence interval |  |  |  |

**Figure 1.**
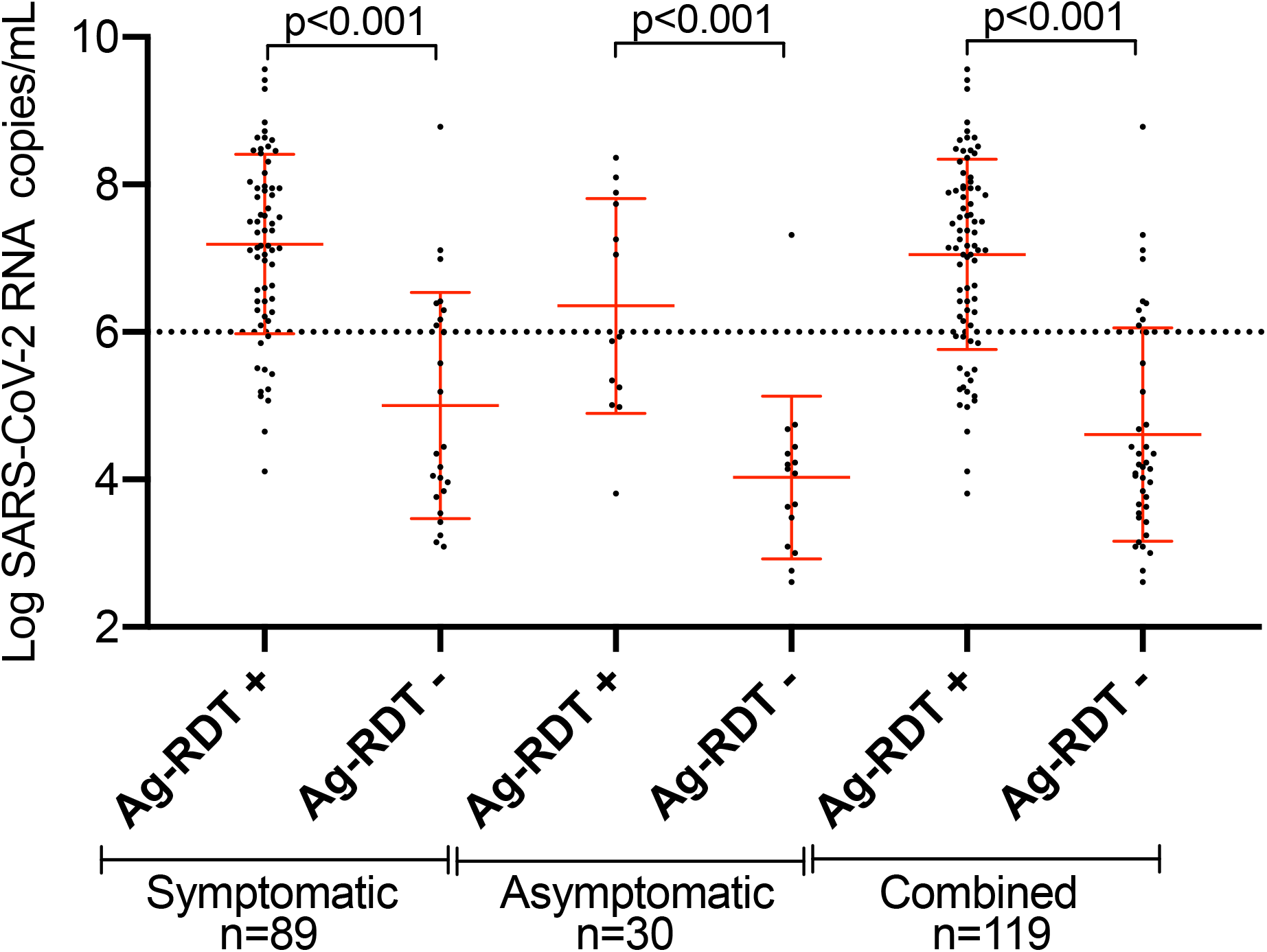
SARS-CoV-2 viral load expressed in log RNA copies/mL among RT-PCR-positive individuals according to Panbio™ RDT results. RDT: antigen-based rapid diagnostic test; RT-PCR reverse transcription polymerase chain reaction; RNA: ribonucleic acid; SARS-CoV-2: severe acute respiratory syndrome coronavirus 2

Sensitivity varied according to RT-PCR VL, even though false-negative results occurred throughout all VL values. Sensitivity was highest at 0.94 in specimens with VL >1.0E7 copies/mL (95%CI 0.87-1.00), decreased slightly to 0.87 (95%CI 0.79-0.95) for specimens >1.0E6 copies/mL, an assumed cut-off for the presence of infectious virus, and to 0.86 (95%CI 0.79-0.93) for specimens >1.0E5 copies/mL. Sensitivity then dropped to 0.76 (95%CI 0.67-0.84) and 0.67 (95%CI 0.59-0.76) for specimens >1.0E4 copies/mL and >1.0E3 copies/mL, respectively (Figure 2).

**Figure 2.**
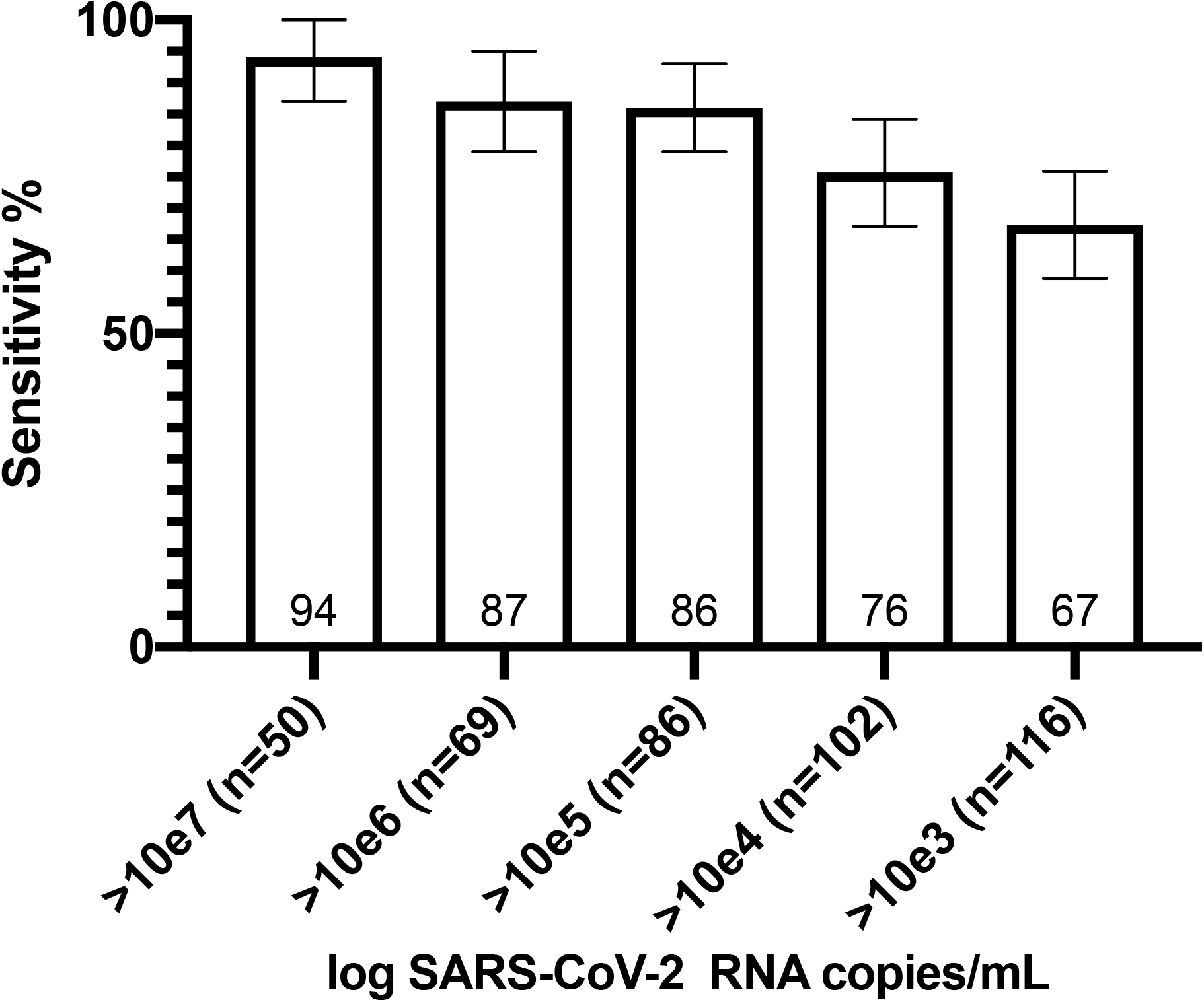
Sensitivity of Panbio™ RDT according to SARS-CoV-2 viral load expressed in log RNA copies/ml. RDT: antigen-based rapid diagnostic test; RNA: ribonucleic acid; SARS-CoV-2: severe acute respiratory syndrome coronavirus 2

Mean viral load was lower children <12 years old than in older children (5.8 [SD 1.8] vs 6.6 [SD 1.7]; p=0.027) and there was a trend towards lower P-RDT sensitivity in children <12 years old (0.57 [95%CI 0.44-0.70]) than in older children (0.74 [95%CI 0.63-0.85]; p=0.057).

### Symptomatic participants

Among the 533 (64.9%) symptomatic participants, median duration of symptoms at time of testing was two days (IQR 1-3) (eTable 2 in the Supplement). The most frequently reported symptoms were headache (56%), nasal discharge (56%), cough (45%) and fatigue (44%) (eTable 2 in the Supplement). Eighty-nine symptomatic patients (16.7%) were positive by RT-PCR with a mean RNA VL of log 6.6 copies/mL (SD 1.6) (Table 1). The P-RDT displayed an overall sensitivity and specificity of 0.73 (95%CI 0.64-0.82) and 1.00 (95%CI 0.99-1.00) respectively (Table 2 and eTable 1 in the Supplement). For specimens with VL >1.0E6 copies/mL, sensitivity was 0.87 (95%CI 0.79-0.95). Mean VL was higher among positive P-RDT specimens than negative ones (7.2 [SD 1.2] vs 5.0 [SD 1.5]; p<0.001) (Figure 1).

Sensitivity was 0.68 at 0-1 DPOS (95%CI 0.53-0.83), peaked at 1.00 at 2 DPOS (95%CI 1.00-1.00), then gradually decreased to 0.73 (95%CI 0.46-0.99), 0.63 (95%CI 0.29-0.96) and 0.56 (95%CI 0.23-0.88) at 3, 4 and 5 DPOS respectively (Figure 3A). False-negative results occurred across all DPOS except at 2 DPOS.

**Figure 3.**
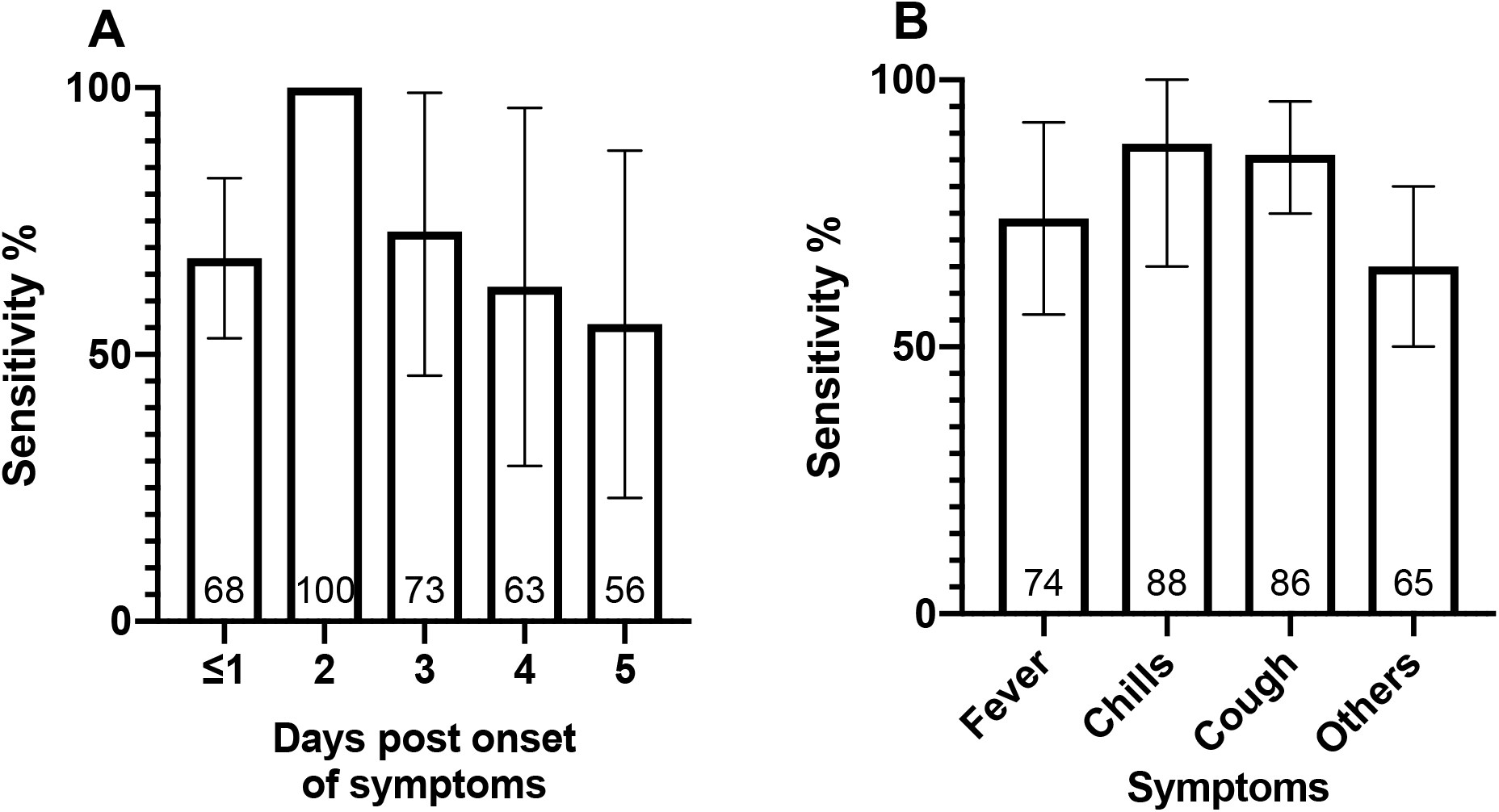
Sensitivity of Panbio™ RDT according to days post onset of symptoms (A) and clinical symptoms (B). RDT: antigen-based rapid diagnostic test

Additionally, we analyzed sensitivity according to typical acute COVID-19 symptoms. Only objective symptoms were reported for the purpose of this analysis, because of the low ability of children to report more subjective symptoms such as anosmia, even though very suggestive of COVID-19.

Sensitivity was highest in the presence of chills (0.88 [95%CI 0.65-1.00]) and cough (0.86 [95%CI 0.75-0.96]), followed by fever (0.74 [95%CI 0.56-0.92]), and then non-specific symptoms (0.65 [95%CI 0.50-0.80]) (Figure 3B).

There was a trend towards lower P-RDT sensitivity in children <12 years old (0.62 [95%CI 0.45-0.78]) than in older ones (0.80 [95%CI 0.69-0.91]; p=0.09).

### Asymptomatic participants

Among the 289 (35.1%) asymptomatic participants at time of sampling, 10.4% (30/289) were positive by RT-PCR with a mean RNA VL of log 5.1 copies/mL (SD 1.7), which was significantly lower than found in symptomatic participants (p<0.001) (Table 1). The P-RDT displayed an overall sensitivity and specificity of 0.43 (95%CI 0.26-0.61) and 1.00 (95%CI 1.00-1.00) respectively (Table 2 and eTable 1 in the Supplement). For specimens with VL >1.0E6 copies/mL, sensitivity was 0.86 (95%CI 0.60-1.11). Mean VL was higher among positive P-RDT specimens than in negative ones (6.4 [SD 1.5] vs 4.0 [SD 1.1]; p<0.001) (Figure 1).

## DISCUSSION

This study prospectively evaluated the diagnostic accuracy of the Panbio™-COVID-19 RDT in the clinical setting in more than 800 symptomatic and asymptomatic children, taking into account VL, DPOS, and clinical parameters. The study was performed during a period of sustained virus circulation, with an overall positivity RT-PCR rate of 17% among symptomatic participants. The major finding of our work is an overall suboptimal 66% sensitivity of the assay, respectively ranging between 43% and 73% in asymptomatic and symptomatic children. On the other hand, specificity was 100% regardless of the presence or absence of symptoms. It would therefore seem very unlikely that children are unnecessarily sent into quarantine, which is important from a public health perspective. The WHO RDT target product profile cut-off of ≥80% for sensitivity and ≥97% for specificity, was not achieved in terms of sensitivity^13^. The relatively low sensitivity of the P-RDT is in line with previous data showing an assay sensitivity of 45-78% among symptomatic children^5,8,9^, and confirms that the assay sensitivity is lower than that in symptomatic adults in whom the largest studies report sensitivity between 80-92%^5-7^. However, among symptomatic children with VL >1.0E6 copies/mL, the assay’s sensitivity (87%) was only marginally lower than symptomatic adults with similar VL (96%)^6^. The suboptimal sensitivity of the assay in children is most likely explained by the increasingly recognized evidence that children have lower SARS-CoV-2 VLs than adults. Indeed, although initial studies suggested similar VLs in adults and children, they were limited in sample size and did not take into account DPOS^12,14,15^, which is a major determinant of VL^16^. Recently, studies on larger datasets and/or taking into account DPOS have shown that SARS-CoV-2 infected children have significantly lower VLs than adults(^17^ and Bellon, L’Huillier & Eckerle [in revision]). Another possible though unlikely explanation for the lower sensitivity could be the technical challenge of the NPS procedure in children. However, it is unclear how sensitivity would be impacted as this would also affect the yield of RT-PCR. Novel findings in our study relate to the evaluation of P-RDT sensitivity in the light of several factors such as VL, DPOS and clinical symptoms, which has not been reported so far in the pediatric population. As expected, VL was higher among those with positive P-RDT (true positives) than among those with negative P-RDT (false negatives), as already shown in the adult setting^6^. Similarly, and as previously shown in adults^2,3,6^, sensitivity was correlated with VL, peaking at 94% in specimens with >1.0E7 copies/mL and dropping to 67% in specimens >1.0E3 copies/mL. A respective 86% and 87% sensitivity remained good in specimens >1.0E5 copies/mL and >1.0E6 copies/mL (the accepted cut-off for the presence of infectious virus^11,16^). However, the presence of false-negative P-RDT results in participants with VLs compatible with shedding of infectious virus is important from a public health perspective as they would not be identified by P-RDT despite being likely contagious. Another interesting finding was the impact of DPOS on the assay sensitivity. Sensitivity was optimal at 2 DPOS, when VL is expected to peak ^18,19^. These findings somehow differ from adult data where sensitivity remained high throughout the first five DPOS (even though lower at 0 DPOS)^6^, likely here again reflecting the impact of higher VLs in adults on P-RDT sensitivity. Similarly, the trend towards lower sensitivity in children <12 years old is likely explained by lower VLs in younger children, as seen in this study and other publications (^17^ and Bellon, L’Huillier & Eckerle [in revision]). Among symptomatic participants, sensitivity was better in those with COVID-19 typical symptoms, as previously shown in adults^6^. Interestingly, the sensitivity of P-RDT in children asymptomatic at time of sampling was low at 43%. This is probably related to the fact that asymptomatic children in our dataset had significantly lower VL than symptomatic children.

The strength of our study is related to the large size of a pure pediatric dataset. The large subset of asymptomatic children at time of sampling, and the analysis of diagnostic accuracy based on VL, DPOS and specific symptoms represent additional strengths and novelties. With a majority of mild clinical presentation and 25% of asymptomatic among RT-PCR positive cases, our dataset is representative of the majority of SARS-CoV-2 infected children, even though the extent of the pediatric contribution to community transmission is still debated.

Our study has several limitations. First, the evaluation was based on one RDT only. Comparative studies have shown similar or reduced performance of other RDTs when compared to the P-RDT^6,20^. Is it therefore highly unlikely that any RDT would perform significantly better in children than the P-RDT. Second, the study was performed using two different validated RT-PCR assays as gold standards, although 90% of the specimens were tested on the Cobas and standard curves used to calculate VL from Ct values were previously validated and used in several publications^6,11,12^. Third, the study was conducted in a high prevalence setting. Extrapolating the findings to low prevalence settings must be done with caution. Then, providing that the NPS for P-RDT was always performed after the NPS for RT-PCR, one cannot exclude that the second procedure was more challenging to perform. Finally, we did not evaluate the performance of P-RDT on oropharyngeal, nasal or saliva specimens. However, given the fact that VL is lower in these anatomical compartments when compared to NPS^21-24^, one can expect even lower sensitivity of P-RDT if used on oropharyngeal, nasal or saliva specimens.

In conclusion, this independent study confirms the respective suboptimal sensitivity of P-RDT in symptomatic children, and its poor sensitivity in asymptomatic children at time of sampling, providing additional evidence for cautious routine use of these tests for the detection of SARS-CoV-2, both in symptomatic and asymptomatic children. This study also highlights the impact of VL, DPOS and clinical presentation on the assay’s sensitivity and show that the sensitivity was ≥80% in participants with VLs >1.0E5 copies/mL, suggesting reliable identification of contagious individuals^11,16^. However, it should be discussed whether missing individuals with lower VLs is acceptable, since they might subsequently have an increase in their VL and become contagious. Therefore, public health benefits of rapidly identifying infected children should be balanced with the disadvantages of missed diagnoses^25^. For individual diagnosis, P-RDT seems a decent alternative to RT-PCR in symptomatic children ≥12 years, especially if tested <5 DPOS based on our findings. For mass pediatric screening, however, such as in school settings or institutions, providing there is no vulnerable contact person, the suboptimal sensitivity of P-RDT is likely outweighed by the advantages of P-RDT, allowing to rapidly identify most infected individuals without the need of a laboratory facility.

## Data Availability

data are available upon reasonable request upon peer-reviewed manuscript publication

## Acknowledgements

We thank our medical students (Mélina Ben Allel, Emmanuelle Mohbat, Liv Mahler, James Nef, Mia Lidén, Eloïse Sattonnay, Estelle Delamare, Eva Dufeil, Mariella Baccaro, Micaela Ruef, Natacha Pougnier, Sami Uslu, Sara Bekiri, Sara Moreira, Sophie Margot, Thomas Demaurex, Vanesa Redzepi) for their help in the recruitment of patients, nurses and staff of the Pediatric Emergency Department of our institution for their work, as well as participants for their willingness to participate in the study.

