## Supplementary Tables and Figure for "Diagnostic accuracy of SARS-CoV-2 rapid antigen detection testing in symptomatic and asymptomatic children in the clinical setting"

**SUPPLEMENT**

**eFigure 1. Study Flowchart**
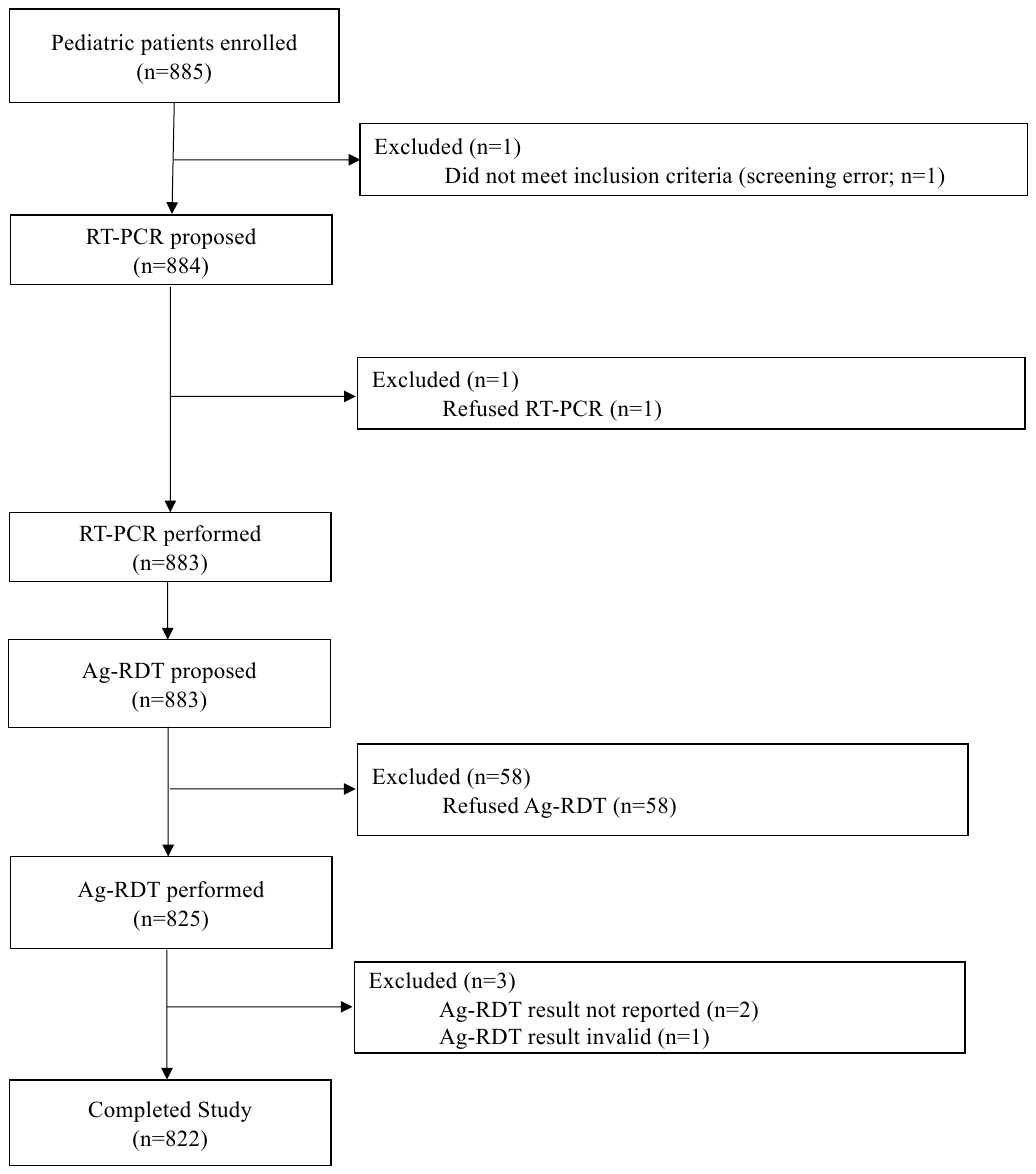

RDT: antigen-based rapid diagnostic test; RT-PCR reverse transcription polymerase chain reaction

**eTable 1. RT-PCR and Panbio^TM^ RDT results**

|  | **Total** | **P-RDT + /**  **RT-PCR +**  **(True positive)** | **P-RDT – /**  **RT-PCR +**  **(False negative)** | **P-RDT + /**  **RT-PCR –**  **(False positive)** | **P-RDT – /**  **RT-PCR –**  **(True negative)** |
| --- | --- | --- | --- | --- | --- |
| Symptomatic | 533 | 65 | 24 | 1 | 443 |
| Asymptomatic | 289 | 13 | 17 | 0 | 259 |
| Combined | 822 | 78 | 41 | 1 | 702 |

RDT: antigen-based rapid diagnostic test; P-RDR: Panbio^TM^ RDT; RT-PCR reverse transcription polymerase chain reaction

**eTable 2. Characteristics of symptomatic study participants**

|  | **Symptomatic**  **(n=533)** |
| --- | --- |
| Symptoms, n (%)  Headache  Nasal Discharge  Cough  Fatigue  Dysphagia  Fever  Abdominal pain  Myalgia  Diarrhea  Reduced intake  Chills  Vomiting  Shortness of breath  Dysgueusia/agueusia  Anosmia  Nausea  Irritability | 297 (55.7)  296 (55.5)  237 (44.5)  235 (44.1)  219 (41.1)  149 (30.0)  106 (19.9)  84 (15.8)  79 (14.8)  77 (14.4)  45 (8.4)  43 (8.1)  42 (7.9)  36 (6.8)  35 (6.6)  25 (4.7)  10 (1.9) |
| Median DPOS to RT-PCR (±IQR) | 2.0 (1.0-3.0) |

RT-PCR: reverse transcription polymerase chain reaction;

DPOS: days post onset of symptoms; IQR: interquartile range
